# National Incidence, Outcomes, and Management Strategies for Pre- and Post-Transplant Atrial Fibrillation in Heart Transplant Recipients

**DOI:** 10.1101/2024.05.14.24307382

**Authors:** Alexander K Karius, Alice L Zhou, Jessica M. Ruck, Allan B. Massie, Dorry L. Segev, David Spragg, Ahmet Kilic

## Abstract

**Background:** Among heart transplant candidates, atrial fibrillation (AF) is a common comorbidity; however, little is known about the impact of pre-transplant AF on incidence of post-transplant AF or other transplant outcomes.

**Methods:** Adult heart transplant recipients transplanted from 07/01/2012 to 07/01/2021 with data available in both the Scientific Registry of Transplant Recipients and Symphony Health pharmacy databases were included. Recipients were categorized by presence of pre-transplant AF using prescription fill data. Perioperative outcomes and survival out to 5 years post-transplant were compared between those with and without pre-transplant AF.

**Results:** Of the 11,789 heart transplant recipients, 2,477 (21.0%) had pre-transplant AF. Pre-transplant AF was associated with an increased likelihood of pre-discharge stroke (aOR 2.13 [95%CI: 1.07-4.26], p=0.03) and dialysis (aOR 1.45 [1.05-2.00], p=0.02), as well as of post-transplant AF at 6 months (aOR 2.42 [1.44-1.48], p=0.001) and 1 year (aOR 2.81 [1.72-4.56], p<0.001). Pre-transplant AF was associated with increased post-transplant mortality at 30 days (aHR 2.39 [1.29-4.44], p=0.006) and 1 year (aHR 1.46 [95% CI: 1.01-2.13], p=0.04), but similar mortality at 5 years (aHR 1.23 [0.96-1.58], p=0.11).

**Conclusion:** Heart transplant recipients with pre-transplant AF had worse short-term outcomes and increased risk of developing post-transplant AF but comparable survival at 5 years post-transplant. Our findings emphasize the importance of increased monitoring for perioperative complications and highlight the long-term safety of heart transplantation in this population.

**What Is New?:**

- Patients with atrial fibrillation who undergo heart transplantation have worse short term survival (30-days and 1-year) but similar long term survival (5-years) compared to recipients without pre-transplant atrial fibrillation.
- Pre-transplant atrial fibrillation increases the risk of clinically significant post-transplant atrial fibrillation and peri-operative stroke.
- Rate vs rhythm control pharmacotherapy for atrial fibrillation is not associated with differences in survival in heart transplant recipients with pre-transplant atrial fibrillation

**What are the Clinical Implications?:**

- Atrial fibrillation should not deter heart transplantation in appropriate candidates, though cardiovascular and stroke risk adjustment may be warranted.
- Use of amiodarone at doses ≤ 200 mg/day is not associated with reduced survival in heart transplant recipients with pre-transplant atrial fibrillation.

## INTRODUCTION

Atrial fibrillation (AF) is the most common clinical arrhythmia, with an estimated prevalence of approximately 13% in adults over the age of 65.^1^ AF is even more common in patients with advanced heart failure, making it an important comorbidity to consider in heart transplant recipients.^2–4^ Despite the fact that AF is a highly prevalent and clinically significant comorbidity in the pre- and post-transplant setting, there is a scarcity of literature on the impacts of AF on recipient outcomes..^5^

To our knowledge, published data on the association between pre-transplant AF and post-transplant outcomes is limited to a single-center study of 639 heart transplant recipients in Germany.^6^ The authors reported that pre-transplant AF was associated with increased 1-year post-transplant mortality in heart transplant recipients, but they did not assess any longer-term outcomes. The development of post-transplant AF following heart transplant has been more widely reported. However, incidence rates in current literature vary widely, ranging between 0.3-14%.^5,7–9^ This may be due to the relatively high rate of self-resolving atrial arrythmias that arise in the post-transplant period that are not clinically relevant and do not require treatment.^5^ Additionally, selection of appropriate pharmacotherapy for AF in the heart transplant candidate remains controversial.^10^ However, there is a current lack of studies that investigate the potential impact of pre-transplant amiodarone use at typical AF doses on post-transplant outcomes and whether rate-vs. rhythm-control strategies are associated with differences in post-transplant outcomes.

Given the high estimated prevalence of AF in patients undergoing orthotopic heart transplant and the lack of population level data on associated outcomes, further data is needed on the impacts of pre-transplant AF on post-transplant outcomes. We reviewed national data on recipients of orthotopic heart transplants between 2012 and 2021 to investigate whether the presence of pre-transplant AF is associated with differences in post-transplant outcomes, whether the development of clinically significant AF post-transplant is associated with differences in long-term survival, and whether pharmacologic rate or rhythm control for patients with pre-transplant AF is associated with a difference in post-transplant survival.

## METHODS

### Data source

This study used data from the Scientific Registry of Transplant Recipients (SRTR). The SRTR data system includes data on all donor, wait-listed candidates, and transplant recipients in the U.S., submitted by the members of the Organ Procurement and Transplantation Network (OPTN). The Health Resources and Services Administration (HRSA), U.S. Department of Health and Human Services provides oversight to the activities of the OPTN and SRTR contractors. This dataset has previously been described elsewhere.^11^

### Study population and definitions

We included all adult (≥18 years) heart transplant recipients transplanted between 07/01/2012 and 07/01/2021 in the SRTR database who also had data available in the Symphony Health pharmacy claims database (81.4% capture). We excluded heart-lung transplants, re-transplants, and recipients bridged with durable left ventricular assist device (LVAD).

We categorized recipients by whether or not they had pre-transplant AF. Because the SRTR database does not specifically identify patients with AF, we utilized pre-transplant prescription fill data to identify patients with comorbid AF. We classified recipients who filled a prescription for either (1) a rhythm control agent (amiodarone of 200mg/day or less, dronedarone, flecainide, dofetilide, or sotalol); or (2) a rate control agent (carvedilol, metoprolol, labetalol, or diltiazem) and anticoagulation (apixaban, betrixaban, bivalirudin, dabigatran, dalteparin, danaparoid sodium, desirudin, edoxaban, enoxaparin, fondaparinux, rivaroxaban, tinzaparin, or warfarin) during the 6 months pre-transplant as having AF. We specified the amiodarone dose (≤200mg/day) to reflect typical dosages prescribed for management of AF, as opposed to ventricular arrhythmias. We excluded recipients bridged with durable LVAD given that many of these recipients would be on both a rate-control agent and anticoagulation pre-transplant, and we wanted to avoid misclassifying these recipients as having pre-transplant AF. Recipients were followed until the outcome of interest or administrative censoring on 02/01/2022. This study was approved by the Johns Hopkins Institutional Review Board (IRB00392902).

### Baseline characteristics

We compared baseline donor and recipient characteristics between recipients with and without pre-transplant AF. We assessed normality of all variables using Shapiro-Wilk testing. We reported normally-distributed continuous variables as mean (standard deviation), non-normally-distributed continuous variables as median (interquartile range [IQR]), and categorical variables as number (percentage). We compared normally-distributed continuous variables using Student’s t-tests, non-normally-distributed continuous variables using Wilcoxon rank-sum tests, and categorical variables using Chi-squared tests.

### Perioperative outcomes

We compared pre-discharge acute rejection, stroke, dialysis, and need for a permanent pacemaker, as well as AF within 6 months and 1 year post-transplant using Chi-squared tests and multivariable logistic regression between recipients with and without pre-transplant AF. We compared hospital length of stay (LOS) using Wilcoxon rank-sum tests. Multivariable models were adjusted for variables chosen *a priori,* including donor age, sex, and race; recipient age, sex, race, diagnosis, body mass index (BMI), hypertension, creatinine, and pre-transplant mechanical circulatory support (MCS) (including intra-aortic balloon pump [IABP], temporary ventricular assist devices [tVADs], and extracorporeal membrane oxygenation [ECMO]); and ischemic time.

### Post-transplant survival

We used time-to-event analysis to assess survival out to 5 years post-transplant and Kaplan Meier curves to visualize the incidence of this outcome. We used Cox regression to compare survival of recipients with and without pre-transplant AF at 30 days, 1 year, and 5 years post-transplant. We compared the post-transplant cause of death within 5 years of recipients with and without pre-transplant AF using Chi-squared testing. Multivariable models were adjusted for variables chosen *a priori,* including donor age, sex, and race; recipient age, sex, race, diagnosis, BMI, hypertension, creatinine, and pre-transplant MCS; and ischemic time.

### Subgroup analysis by control agent

Among recipients with pre-transplant AF, we performed a subgroup analysis of outcomes by pre-transplant AF control agent (rate-only control vs. rhythm-only control vs. rate and rhythm control). We compared baseline characteristics, perioperative outcomes, and post-transplant survival as described above.

### Factors associated with survival in recipients with pre-transplant AF

Among recipients with pre-transplant AF, we examined factors associated with 5-year survival using multivariable Cox regression. We separately used multivariable Cox regression to examine the association between the development of AF within 6 months post-transplant on 5-year mortality in those with pre-transplant AF. We adjusted for donor age, sex, and race; recipient age, sex, race, diagnosis, BMI, hypertension, creatinine, pre-transplant MCS, and pre-transplant amiodarone use; and ischemic time.

### Subgroup analysis by post-transplant AF

Among recipients with at least 6 months follow-up, we investigated risk factors associated with the development of post-transplant AF within 6 months. Among recipients who survived at least 6 months post-transplant, we also compared 5-year survival for recipients with and without AF within 6 months post-transplant using Cox regression, adjusting for donor age, sex, and race; recipient age, sex, race, diagnosis, BMI, hypertension, creatinine, and pre-transplant MCS; and ischemic time. All statistics were performed using StataSE 18 (StataCorp, College Station, Texas).

## RESULTS

### Baseline characteristics

Of a total 11,789 recipients that were included in the study, 2,477 (21.0%) had pre-transplant AF. Donors for transplant recipients with versus without pre-transplant AF were more likely to be male (69.6% vs. 66.9%, p=0.01; **Table 1**). Recipients with versus without pre-transplant AF were more likely to be male (74.1% vs. 68.5%, p<0.001) and of White race (67.6% vs. 63.9%, p<0.001) and less likely to require pre-transplant ECMO (2.5% vs. 3.6%, p=0.01). Among those transplanted in the pre-2018 era, recipients with versus without pre-transplant AF were less likely to be status 1A (56.8% vs. 63.5%, p<0.001). Among those transplanted in the post-2018 era, recipients with and without pre-transplant AF had similar distribution of statuses (p=0.09).

**Table 1:** Baseline donor, recipient, and transplant characteristics by pre-transplant atrial fibrillation.

| Variable, n (%) | No pre-transplant AF<br>(n=9,312) | Pre-transplant AF<br>(n=2,477) | p-value |
| --- | --- | --- | --- |
| **Donor characteristics** |  |  |  |
| Age (years), median (IQR) | 31 (23-40) | 31 (23-40) | 0.83 |
| Male sex | 6,227 (66.9%) | 1,724 (69.6%) | 0.01 |
| White race | 5,860 (62.9%) | 1,591 (64.2%) | 0.23 |
| Cause of death |  |  | 0.25 |
| Anoxia | 3,405 (36.6%) | 946 (38.2%) |  |
| Cerebrovascular/stroke | 1,545 (16.6%) | 376 (15.2%) |  |
| Head trauma | 4,095 (44.0%) | 1,090 (44.0%) |  |
| Other | 264 (2.8%) | 65 (2.6%) |  |
| DCD | 77 (0.8%) | 20 (0.8%) | 0.92 |
| **Recipient characteristics** |  |  |  |
| Age (years), median (IQR) | 57 (47-64) | 57 (48-64) | <0.001 |
| Male sex | 6,379 (68.5%) | 1,835 (74.1%) | <0.001 |
| Race |  |  | <0.001 |
| White | 5,952 (63.9%) | 1,674 (67.6%) |  |
| Black | 2,061 (22.1%) | 517 (20.9%) |  |
| Other | 1,299 (13.9%) | 286 (11.5%) |  |
| Diagnosis |  |  | 0.007 |
| Ischemic CM | 2,831 (30.4%) | 752 (30.4%) |  |
| Nonischemic dilated CM | 4,665 (50.1%) | 1,309 (52.8%) |  |
| Restrictive CM | 596 (6.4%) | 112 (4.5%) |  |
| Hypertrophic CM | 401 (4.3%) | 108 (4.4%) |  |
| Congenital | 474 (5.1%) | 113 (4.6%) |  |
| Other | 336 (3.6%) | 83 (3.4%) |  |
| Status |  |  |  |
| Pre-2018 |  |  | <0.001 |
| Status 1A | 3,538 (63.5%) | 840 (56.8%) |  |
| Status 1B | 1,754 (31.5%) | 563 (38.1%) |  |
| Status 2 | 279 (5.0%) | 75 (5.1%) |  |
| Post-2018 |  |  | 0.09 |
| Status 1 | 369 (9.9%) | 74 (7.4%) |  |
| Status 2 | 2,154 (57.6%) | 587 (58.8%) |  |
| Status 3 | 396 (10.6%) | 92 (9.2%) |  |
| Status 4 | 533 (14.2%) | 163 (16.3%) |  |
| Status 5 | 43 (1.1%) | 13 (1.3%) |  |
| Status 6 | 246 (6.6%) | 70 (7.0%) |  |
| Pre-transplant MCS |  |  |  |
| IABP | 2,142 (23.0%) | 539 (21.8%) | 0.19 |
| Temporary VAD | 1,006 (10.8%) | 269 (10.9%) | 0.94 |
| ECMO | 332 (3.6%) | 63 (2.5%) | 0.01 |
| Hypertension | 1,252 (56.4%) | 263 (60.2%) | 0.14 |
| COPD | 126 (6.1%) | 19 (4.5%) | 0.21 |
| Creatinine (mg/dL), median (IQR) | 1.18 (.91-1.5) | 1.2 (.98-1.5) | 0.002 |
| BMI (kg/m<sup>2</sup>), median (IQR) | 26.4 (23.2-30.2) | 27.0 (23.7-30.6) | <0.001 |
| Ischemic time (minutes), median (IQR) | 195 (152-232) | 192 (150-231) | 0.14 |

Abbreviations: AF, atrial fibrillation; BMI, body mass index; CM, cardiomyopathy; COPD, chronic obstructive pulmonary disease; DCD, donation after circulatory death; ECMO, extracorporeal membrane oxygenation; IABP, intra-aortic balloon pump; IQR, interquartile range; MCS, mechanical circulatory support; VAD, ventricular assist device.

### Perioperative outcomes

Recipients with pre-transplant AF had greater likelihood of experiencing pre-discharge stroke (aOR 2.13 [95% CI: 1.07-4.26], p=0.03) and pre-discharge dialysis (aOR 1.45 [95% CI: 1.05-2.00], p=0.02; **Table 2**). However, recipients with and without pre-transplant AF had similar likelihood of experiencing acute rejection (aOR 1.06 [0.81-1.40], p=0.67) and requirement for pacemaker (aOR 1.12 [0.67-1.88], p=0.66), as well as similar median hospital lengths of stay (15 [11-23] vs. 16 [11-24] days, p=0.30).

**Table 2:** Unadjusted and adjusted perioperative outcomes by pre-transplant AF.

| **Outcome** | **Unadjusted outcomes** |  |  | **Adjusted outcomes** |  |
| --- | --- | --- | --- | --- | --- |
|  | **No AF**<br>**(n=9,312)** | **AF**<br>**(n=2,477)** | **p-value** | **Adjusted OR** | **Adjusted**<br>**p-value** |
| Acute rejection | 1,806 (19.4%) | 479 (19.3%) | 0.92 | 1.06 (0.81-1.40) | 0.67 |
| Stroke | 219 (2.4%) | 58 (2.4%) | 0.96 | **2.13 (1.07-4.26)** | **0.03** |
| Dialysis | 1,218 (13.2%) | 313 (12.7%) | 0.51 | **1.45 (1.05-2.00)** | **0.02** |
| Pacemaker | 215 (2.3%) | 67 (2.7%) | 0.26 | 1.12 (0.67-1.88) | 0.66 |
| Hospital LOS | 16 (11-24) | 15 (11-23) | 0.30 |  |  |
| AF 6m post-tx | **233 (2.7%)** | **136 (6.0%)** | **<0.001** | **2.42 (1.44-4.08)** | **0.001** |
| AF 1y post-tx | **244 (3.2%)** | **151 (7.4%)** | **<0.001** | **2.81 (1.72-4.56)** | **<0.001** |

Abbreviations: HR, hazards ratio; LOS, length of stay
Models adjusted for donor age, sex, race; recipient age, sex, race, diagnosis, BMI, hypertension, creatinine, IABP, tVAD, ECMO, ischemic time

Overall incidence of AF within 6 months post-transplant was 3.4% (n=369) and within 12 months post-transplant was 4.1% (n=395). Pre-transplant AF was associated with greater risk for post-transplant AF at 6 months (aOR 2.42 [95% CI: 1.44-1.48], p=0.001) and 1 year (aOR 2.81 [1.72-4.56], p<0.001) post-transplant (**Table 2**).

### Post-transplant survival

Pre-transplant AF was associated with greater risk for mortality at 30 days (aHR 2.39 [95% CI: 1.29-4.44], p=0.006) and 1 year post-transplant (aHR 1.46 [95% CI: 1.01-2.13], p=0.04), but similar risk for mortality at 5 years post-transplant (aHR 1.23 [95% CI: 0.96-1.58], p=0.11; **Figure 1**). There were no differences in primary causes of death within 5 years between those with and without pre-transplant AF (**Supplemental Table 1**).

**Figure 1:**
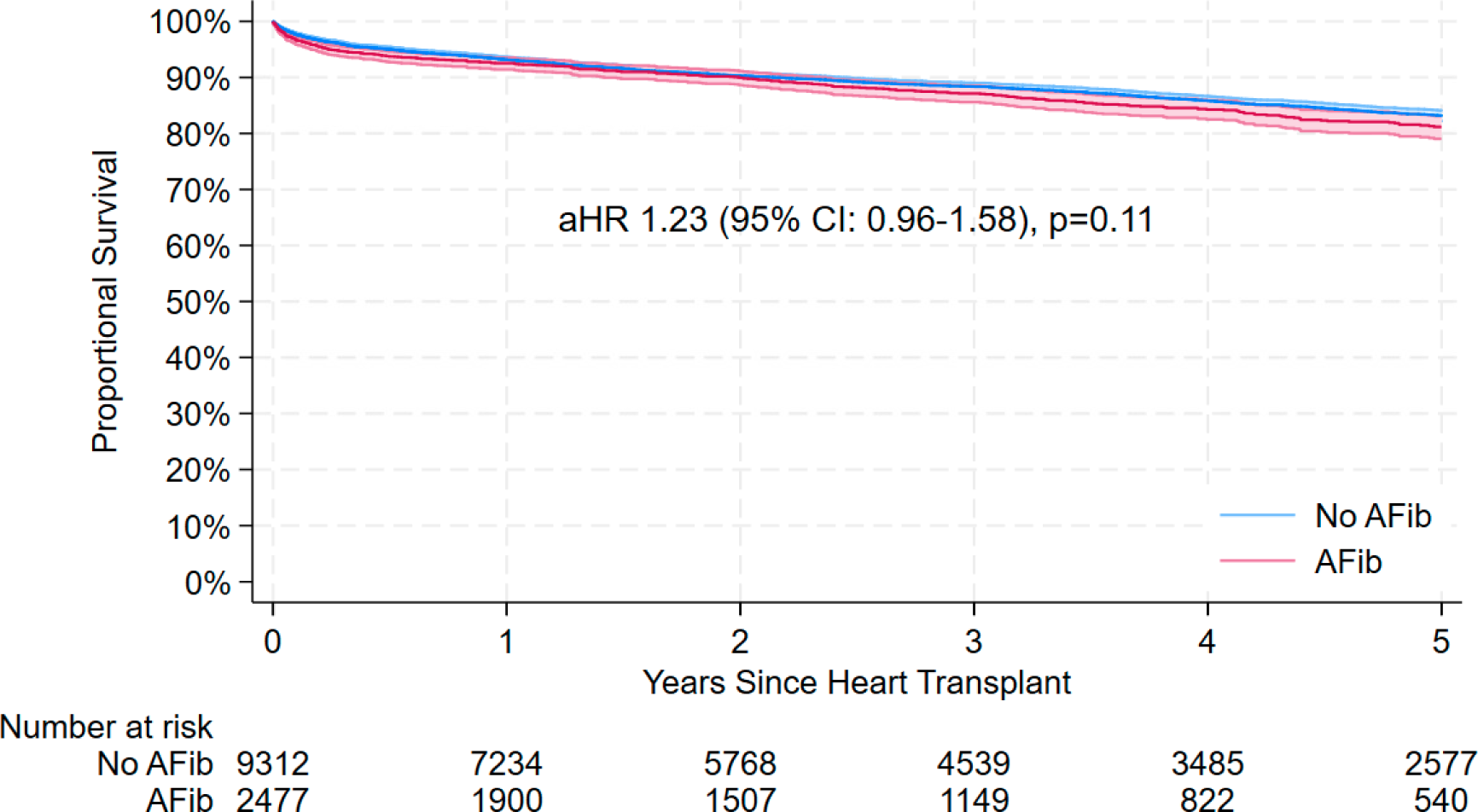
Unadjusted 5-year post-transplant survival by pre-transplant atrial fibrillation.

### Subgroup analysis by control agent

In recipients with pre-transplant AF, we performed a subgroup analysis by AF pharmacologic control agent (rate-only control vs. rhythm-only control vs. rate and rhythm control). Of the 2,477 recipients with pre-transplant AF, 1,315 (53.1%) were on rate control agents only, 609 (24.6%) were on rhythm control agents only, and 553 (22.3%) were on both rate and rhythm control agents. Baseline characteristics for these groups are shown in **Supplemental Table 2**. The breakdown of the specific rhythm control agents utilized is shown in **Supplemental Table 3**. The most common rhythm control agent utilized was amiodarone (68.6%). There were no differences in likelihood of acute rejection, stroke, dialysis, pacemaker insertion, or development of AF within 6 months or 1 year post-transplant (**Supplemental Tables 4 and 5**). There were no differences in adjusted survival between recipients that were on rhythm-only or rate and rhythm control agents compared to rate-only agents (**Table 3**).

**Table 3:**
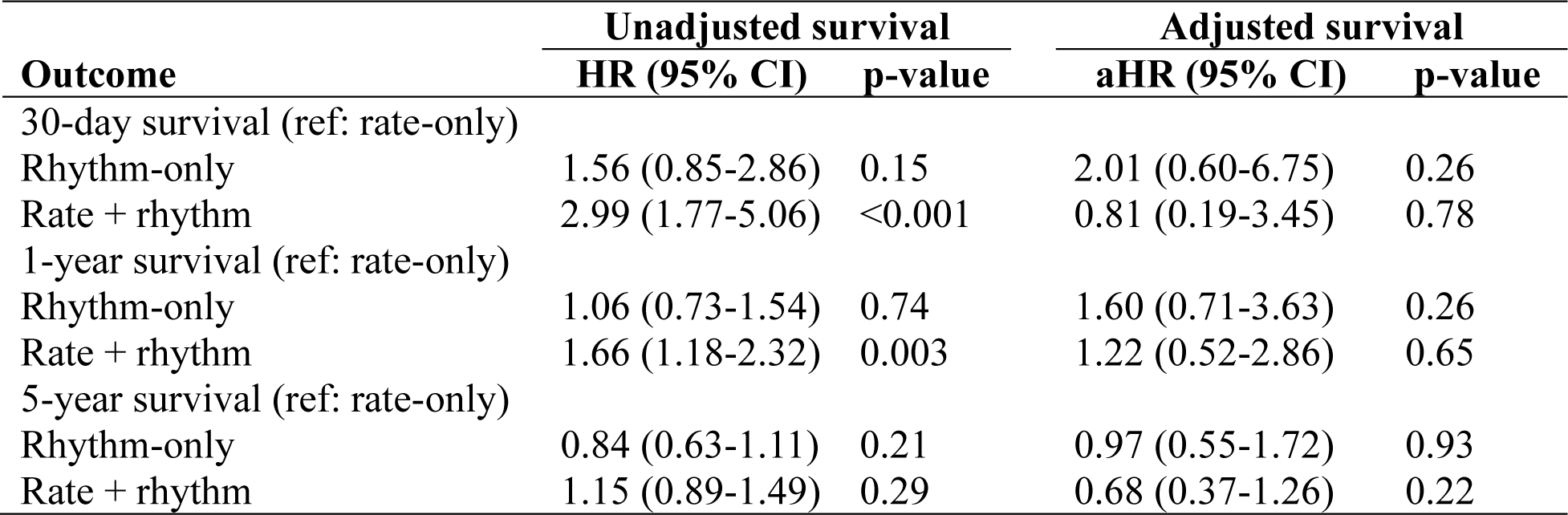
Unadjusted and adjusted survival among recipients with pre-transplant AF, by pre-transplant control agent.

### Factors associated with survival in recipients with pre-transplant AF

Higher recipient creatinine was associated with increased 5-year mortality in those with pre-transplant AF (aHR 1.27 [95% CI: 1.10-1.47], p=0.001), while donor white race was associated with decreased 5-year mortality (aHR 0.54 [95% CI: 0.34-0.86], p=0.009; **Table 4**). Pre-transplant amiodarone compared to other pharmacological control agents was not associated with increased 5-year mortality in those with pre-transplant AF (aHR 0.87 [95% CI: 0.52-1.45], p=0.59). In a subgroup analysis of recipients with pre-transplant AF who survived to 6 months post-transplant, post-transplant AF was not associated with 5-year mortality in those with pre-transplant AF on adjusted (aHR 0.39 [95% CI: 0.05-2.86], p=0.35) analysis.

**Table 4:** Risk factors associated with 5-year mortality in those with pre-transplant atrial fibrillation.

| Variable | Adjusted Hazards Ratio | 95% Confidence Interval | p-value |
| --- | --- | --- | --- |
| **Donor characteristics** |  |  |  |
| Age (per year) | 1.01 | 0.99-1.03 | 0.32 |
| Male sex | 1.16 | 0.67-2.01 | 0.59 |
| White race | 0.54 | 0.34-0.86 | 0.009 |
| **Recipient characteristics** |  |  |  |
| Age (per year) | 1.00 | 0.98-1.03 | 0.77 |
| Male sex | 0.70 | 0.40-1.22 | 0.21 |
| Race (ref: White) |  |  |  |
| Black | 1.11 | 0.61-2.02 | 0.73 |
| Other | 1.40 | 0.67-2.92 | 0.38 |
| Diagnosis (ref: ischemic CM) |  |  |  |
| Nonischemic dilated CM | 0.88 | 0.52-1.48 | 0.62 |
| Restrictive CM | 0.71 | 0.16-3.09 | 0.65 |
| Hypertrophic CM | 0.86 | 0.25-2.92 | 0.81 |
| Congenital | 1.16 | 0.29-4.67 | 0.83 |
| Pre-transplant IABP | 1.30 | 0.59-2.86 | 0.51 |
| Pre-transplant temporary VAD | 0.85 | 0.34-2.18 | 0.74 |
| Hypertension | 1.50 | 0.90-2.50 | 0.12 |
| Creatinine (per mg/dL) | 1.27 | 1.10-1.47 | 0.001 |
| BMI (per kg/m<sup>2</sup>) | 1.00 | 0.99-1.01 | 0.46 |
| Ischemic time (per hour) | 1.12 | 0.91-1.38 | 0.29 |
| Pre-transplant amiodarone | 0.87 | 0.52-1.45 | 0.59 |

Abbreviations: BMI, body mass index; CM, cardiomyopathy; IABP, intra-aortic balloon pump; VAD, ventricular assist device.

### Subgroup analysis by post-transplant AF

Of the 10,853 recipients with at least 6 months of follow-up, 345 (3.2%) had AF during the 6 months post-transplant. Risk factors associated with the development of post-transplant AF in those who survived to 6 months post-transplant were ischemic cardiomyopathy (vs non-ischemic cardiomyopathy) (aOR 2.08 [95% CI: 1.17-3.72], p=0.01), pre-transplant IABP (aOR 2.08 [95% CI: 1.08-4.00], p=0.03), pre-transplant ECMO (aOR 6.23 [95% CI: 1.26-30.76], p=0.03), pre-transplant tVAD (aOR 2.46 [95% CI: 1.29-4.68], p=0.006; **Supplemental Table 6**), and pre-transplant AF (aOR 2.37 [95% CI: 1.38-4.09], p=0.002). Post-transplant AF within 6 months was not associated with adjusted 5-year mortality (aHR 0.35 [95% CI: 0.11-1.09], p=0.07) analysis (**Figure 2**).

**Figure 2:**
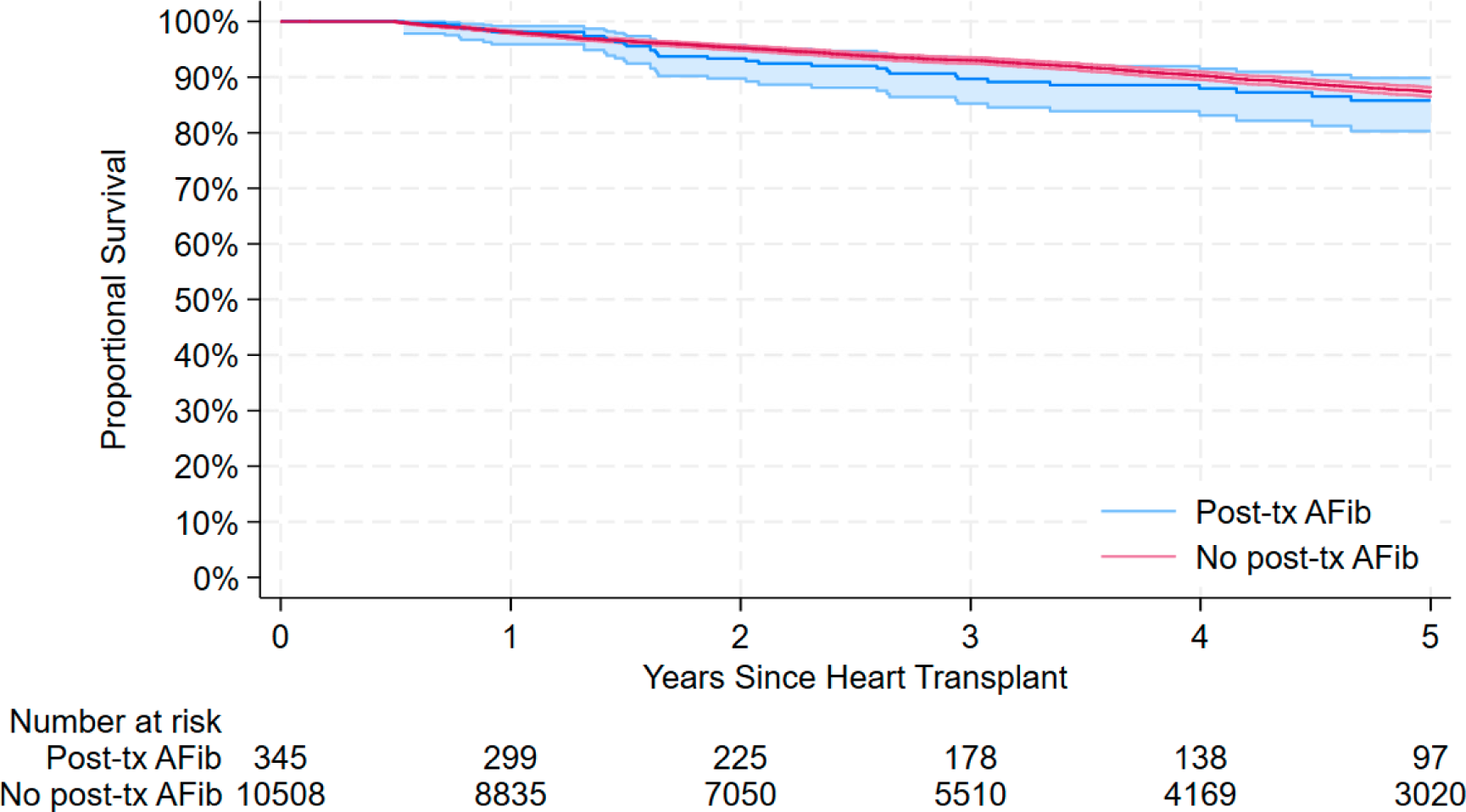
Unadjusted 5-year post-transplant survival conditional on survival at 6 months by atrial fibrillation within 6 months post-transplant.

## DISCUSSION

In this national retrospective analysis, we had a 21% overall pre-heart transplant prevalence of atrial fibrillation. We found that heart transplant recipients with pre-transplant AF had worse short-term survival but similar long-term survival when compared to recipients without pre-transplant AF. We also found that recipients with pre-transplant AF were more likely to experience stroke, require dialysis, and develop AF immediately post-transplant compared to recipients without pre-transplant AF. Further, we found that development of clinically-managed post-transplant AF was not significantly associated with reduced long-term survival among heart transplant recipients, both overall and among those with pre-transplant AF. Lastly, we showed that for recipients with pre-transplant AF, 5-year survival and all studied secondary outcomes were not affected by choice of pre-transplant pharmacotherapy with rate or rhythm control agents.

### Impact of pre-transplant AF on survival

Our finding of increased 1-year post-transplant mortality in patients with pre-transplant AF was consistent with the prior single-center study that reported 1-year post-transplant outcomes from 639 heart transplant recipients at their institution.^6^ The authors reported an absolute difference in 1 year survival between recipients with and without pre-transplant AF of 72.9% vs 80.4% (p=0.04), respectively. However, we were also able to extend our analysis to 5 years post-transplant but noted that, at this timepoint, recipients with pre-transplant AF have similar mortality risk as recipients without pre-transplant AF. We hypothesize that the decreased short-term survival in recipients with pre-transplant AF could potentially be due to perioperative factors related to management of anticoagulation status and the observed increased rate of perioperative stroke. We also analyzed factors associated with post-transplant survival in recipients with pre-transplant AF and found that higher baseline creatinine negatively and white donor race positively associate with five-year survival. Interestingly, development of clinically significant post-transplant AF, in those with pre-transplant AF, did not predict 6-month conditional survival at 5 years post-transplant. These results underscore the long-term safety of transplanting patients with AF, while suggesting a role for more aggressive, early cardiovascular risk adjustment in this population.

### Impact of pre-transplant AF on secondary outcomes

No prior studies from the modern era have directly reported on the association of pre-transplant AF and secondary outcomes such as stroke or acute rejection in heart transplant recipients. This is especially important as proper pharmacotherapy management, especially regarding anticoagulation, is not well understood.^12^ Here we report that pre-transplant AF is significantly associated with an increased risk for stroke, dialysis requirement, and the development of post-transplant AF on adjusted analysis. The increased risk for perioperative stroke may be due to thromboembolic complications of AF, as the higher observed rate of post-transplant AF by 6 months post-transplant suggests that this population may also experience a higher AF burden in the peri-operative period. Alternatively, hemorrhagic complications related to anticoagulant use in this population may contribute to higher rates of perioperative stroke. Pre-transplant AF was not, however, associated with increased risk of acute rejection or post-transplant pacemaker implantation, or with increased post-transplant hospital length of stay. These findings highlight the importance of careful perioperative management and stroke prevention in heart transplant patients with pre-transplant AF considering the increased short-term mortality in this population.

### Incidence and outcomes associated with post-transplant AF

Incidence of AF following orthotopic heart transplant is difficult to determine as many patients experience transient, self-resolving arrythmias for which the diagnosis and classification of is unclear. This has likely contributed to broad reports of post-transplant AF incidence. Our findings of 3.4% 6-month and 4.1% 1-year incidence of pharmacologically managed AF for all transplant recipients is within the range of reported rates in literature of 0.3-14%.^5,7–9^ Given the transient nature of many cases of post-transplant AF, our reported incidence rate likely better reflects the clinically significant disease burden of AF in the heart transplant recipient population compared to studies with higher incidence rates. Upon further analysis, we demonstrated that the development of AF in the first 6 months post-transplant does not impact 5-year survival conditional on survival to 6 months. This was true in both the general heart transplant recipient population and for those with pre-transplant AF. Our results contradict a 2018 meta-analysis that reported increased mortality from data pooled from 5 retrospective studies with a total of 1,230 transplant recipients.^13^ The discrepancy in results may be due to a number of factors. The largest included study (with 530 patients) utilized a combined cohort of subtotal and complete heart transplants wherein total orthotopic heart transplant recipients accounted for a minority of the cohort.^14^ Further, subtotal transplant was found to be a significant risk factor for AF development, indicating that incidence of post-transplant AF following total orthotopic heart transplant was likely lower than the reported aggregate value. Additionally, three out of four of the remaining studies combined atrial flutter and AF,^5,15,16^ while the remaining study relied on an outdated patient cohort (88 recipients between 1985-1994).^17^ Lastly, we are specifically reporting on 5-year survival in recipients who survived at least 6 months post-transplant. Taken together, our data demonstrated that, regardless of pre-transplant AF status, recipients with post-transplant AF who survive to 6 months post-transplant have similar long-term survival as recipients who do not develop AF by 6 months. This may be used to guide clinicians in counseling recipients who develop AF following heart transplant.

### Impact of AF pharmacotherapy selection on post-transplant survival

We performed subgroup analyses to assess whether pharmacotherapy with rate or rhythm control agents was associated with any relative benefit to patients with pre-transplant AF undergoing heart transplant. We found no significant differences in survival or secondary outcomes between rate or rhythm control strategies. Further, we specifically demonstrated that use of amiodarone at typical doses for AF is not associated with worse post-transplant survival. This is important as prior literature is divided on whether pre-transplant use of amiodarone worsens post-transplant outcomes, with some studies reporting that it significantly decreases post-transplant survival and others showing no difference.^10,18–21^ Importantly, literature suggests that amiodarone dose may be a significant factor in whether pre-transplant use negatively affects post-transplant outcomes.^22^ However, no prior studies examining the effects of amiodarone on post-transplant outcomes have utilized dosage thresholds for amiodarone, and thus the effect of pre-transplant amiodarone at typical AF doses has not been well described. A strength of our analysis is that we analyzed patient-level prescription fill data and could thus identify the specific dosage of amiodarone each patient was taking prior to transplant and only included patients taking lower doses of amiodarone (200mg/day or less), which better reflects dosages used for AF than for ventricular arrythmia control.

### Limitations

Our study was limited by its retrospective nature and the data available in the SRTR database. Our methodology of identifying patients with atrial fibrillation relied on filled outpatient prescriptions for common atrial fibrillation pharmacotherapy. However, this may not have captured patients that had been treated by catheter ablation and did not take any rate or rhythm pharmacotherapy in the 6 month pre-transplant period. Similarly, we were unable to capture patients that developed clinically significant post-transplant AF but were treated by primary catheter ablation without any standard pharmacotherapy. Since the approach to pharmacotherapy selection is similar for the main two types of AF - paroxysmal and persistent - we were unable to specifically compare outcomes by type of AF. Our inclusion methodology may have also erroneously captured patients taking anticoagulation and rate control agents or rhythm control agents without true AF. We attempted to mitigate this by excluding patients bridged with durable LVAD to transplant, as durable LVAD is an indication for anticoagulation. We also excluded patients taking high dose amiodarone, including only those patients taking typical doses of amiodarone for AF (200 mg/day or less), given that amiodarone for ventricular tachyarrhythmias is commonly dosed at a higher amount. Lastly, our data on survival with the development of post-transplant AF is contingent upon survival to six months, which was necessary so that prescriptions for AF pharmacotherapy could be filled for patients that developed AF. Therefore, the effect of AF on 6-month survivorship was not described.

In conclusion, we demonstrate that pre-transplant AF is associated with increased risk for perioperative stroke, perioperative dialysis, and mortality at 30 days and 1 year post-transplant. However, we also found that pre-transplant AF survival was not associated with increased mortality at 5 years post-transplant. We report a 21% pre-transplant incidence of atrial fibrillation and an overall 3.4% incidence of clinically important post-transplant atrial fibrillation. Lastly, we demonstrate that use of amiodarone at typical atrial fibrillation dosages in heart transplant candidates was not associated with post-transplant outcomes. Overall, our findings underscore the long-term safety of transplanting heart candidates with pre-transplant AF and highlight a potential role for more aggressive, early cardiovascular risk adjustment and increased monitoring of perioperative complications of these patients to improve short-term outcomes.

## Data Availability

All data refer to in the manuscript will be available for any reviewer / future use.

## ACKNOWLEDGEMENTS

This work was supported by the Pozefsky Scholars Program, grant number F32-AG067642091A1 (Ruck) from the National Institute on Aging (NIA), and K24-AI144954-08 (Segev) from The National Institute of Allergy and Infectious Disease (NIAID). The analyses described here are the responsibility of the authors alone and do not necessarily reflect the views or policies of the Department of Health and Human Services, nor does mention of trade names, commercial products or organizations imply endorsement by the U.S. Government.

The data reported here have been supplied by the Hennepin Healthcare Research Institute (HHRI) as the contractor for the Scientific Registry of Transplant Recipients (SRTR). The interpretation and reporting of these data are the responsibility of the author(s) and in no way should be seen as an official policy of or interpretation by the SRTR or the U.S. Government.

## Supplemental Material: Intended for publication as an online data supplement

**Supplemental Table 1:** Causes of death within 5 years in recipients by pre-transplant AF.

| **Variable, n (%)** | **No pre-transplant AF**<br>**(n=1,148)** | **Pre-transplant**<br>**AF**<br>**(n=332)** | **p-value** |
| --- | --- | --- | --- |
| **Causes of death** |  |  | 0.86 |
| Graft Failure - other | 69 (6.0%) | 22 (6.6%) |  |
| Graft Failure - Rejection | 61 (5.3%) | 20 (6.0%) |  |
| Infection | 222 (19.3%) | 67 (20.2%) |  |
| Cardiovascular | 179 (15.6%) | 58 (17.5%) |  |
| Pulmonary | 80 (7.0%) | 24 (7.2%) |  |
| Cerebrovascular | 71 (6.2%) | 16 (4.8%) |  |
| Hemorrhage | 216 (18.8%) | 56 (16.9%) |  |
| Malignancy | 82 (7.1%) | 17 (5.1%) |  |
| Renal Failure | 15 (1.3%) | 3 (0.9%) |  |
| Other | 153 (13.3%) | 49 (14.8%) |  |

**Supplemental Table 2:** Baseline donor, recipient, and transplant characteristics in recipients with pre-transplant atrial fibrillation, by control agent.

| Variable, n (%) | Rate-only<br>(n=1,315) | Rhythm-only<br>(n=609) | Rate+Rhythm<br>(n=553) | p-value |
| --- | --- | --- | --- | --- |
| **Donor characteristics** |  |  |  |  |
| Age (years), median (IQR) | 31 (23-39) | 31 (23-40) | 33 (24-41) | 0.10 |
| Male sex | 910 (69.2%) | 426 (70.0%) | 388 (70.2%) | 0.90 |
| White race | 850 (64.6%) | 390 (64.0%) | 351 (63.5%) | 0.89 |
| Cause of death |  |  |  | 0.22 |
| Anoxia | 519 (39.5%) | 224 (36.8%) | 203 (36.7%) |  |
| Cerebrovascular/stroke | 180 (13.7%) | 100 (16.4%) | 96 (17.4%) |  |
| Head trauma | 575 (43.7%) | 273 (44.8%) | 242 (43.8%) |  |
| Other | 41 (3.1%) | 12 (2.0%) | 12 (2.2%) |  |
| DCD | 10 (0.8%) | 4 (0.7%) | 6 (1.1%) | 0.69 |
| **Recipient characteristics** |  |  |  |  |
| Age (years), median (IQR) | 56 (46-64) | 58 (50-65) | 58 (51-65) | <0.001 |
| Male sex | 953 (72.5%) | 449 (73.7%) | 433 (78.3%) | 0.03 |
| Race |  |  |  | 0.04 |
| White | 853 (64.9%) | 435 (71.4%) | 386 (69.8%) |  |
| Black | 301 (22.9%) | 111 (18.2%) | 105 (19.0%) |  |
| Other | 161 (12.2%) | 63 (10.3%) | 62 (11.2%) |  |
| Diagnosis |  |  |  | 0.58 |
| Ischemic CM | 390 (29.7%) | 188 (30.9%) | 174 (31.5%) |  |
| Nonischemic dilated CM | 711 (54.1%) | 304 (49.9%) | 294 (53.2%) |  |
| Restrictive CM | 62 (4.7%) | 31 (5.1%) | 19 (3.4%) |  |
| Hypertrophic CM | 50 (3.8%) | 30 (4.9%) | 28 (5.1%) |  |
| Congenital | 59 (4.5%) | 34 (5.6%) | 20 (3.6%) |  |
| Other | 43 (3.3%) | 22 (3.6%) | 18 (3.3%) |  |
| Status |  |  |  |  |
| Pre-2018 |  |  |  | 0.32 |
| Status 1A | 475 (58.6%) | 178 (52.0%) | 187 (57.4%) |  |
| Status 1B | 298 (36.8%) | 143 (41.8%) | 122 (37.4%) |  |
| Status 2 | 37 (4.6%) | 21 (6.1%) | 17 (5.2%) |  |
| Post-2018 |  |  |  | 0.83 |
| Status 1 | 42 (8.3%) | 15 (5.6%) | 17 (7.5%) |  |
| Status 2 | 300 (59.4%) | 160 (59.9%) | 127 (55.9%) |  |
| Status 3 | 44 (8.7%) | 22 (8.2%) | 26 (11.5%) |  |
| Status 4 | 80 (15.8%) | 43 (16.1%) | 40 (17.6%) |  |
| Status 5 | 7 (1.4%) | 4 (1.5%) | 2 (0.9%) |  |
| Status 6 | 32 (6.3%) | 23 (8.6%) | 15 (6.6%) |  |
| Pre-transplant MCS |  |  |  |  |
| IABP | 291 (22.1%) | 135 (22.2%) | 113 (20.4%) | 0.69 |
| Temporary VAD | 160 (12.2%) | 44 (7.2%) | 65 (11.8%) | 0.004 |
| ECMO | 39 (3.0%) | 12 (2.0%) | 12 (2.2%) | 0.36 |
| Hypertension | 133 (57.6%) | 61 (59.8%) | 69 (66.3%) | 0.32 |
| COPD | 8 (3.6%) | 5 (5.2%) | 6 (6.1%) | 0.58 |
| Creatinine (mg/dL), median (IQR) | 1.16 (.95-1.5) | 1.2 (1-1.5) | 1.27 (1.02-1.54) | <0.001 |
| BMI (kg/m<sup>2</sup>), median (IQR) | 27.0 (23.7-30.5) | 26.9 (23.1-30.4) | 27.1 (24.3-30.9) | 0.10 |
| Ischemic time (minutes), median (IQR) | 195 (153-230) | 191 (148.5-232) | 188 (148-230) | 0.40 |

Abbreviations: AF, atrial fibrillation; BMI, body mass index; CM, cardiomyopathy; COPD, chronic obstructive pulmonary disease; DCD, donation after circulatory death; ECMO, extracorporeal membrane oxygenation; IABP, intra-aortic balloon pump; IQR, interquartile range; MCS, mechanical circulatory support; VAD, ventricular assist device.

**Supplemental Table 3:**
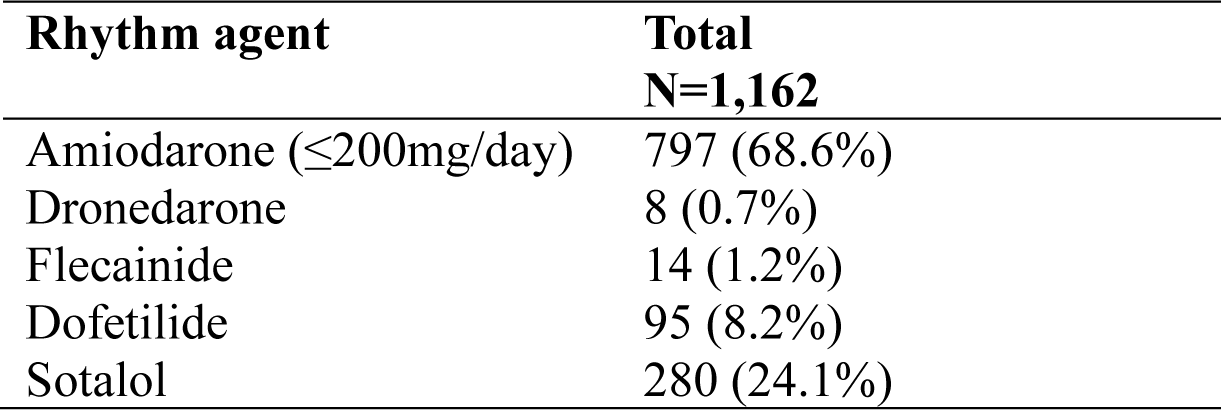
Breakdown of rhythm agents utilized in those with pre-transplant rhythm control.

| **Rhythm agent** | **Total**<br>**N=1,162** |
| --- | --- |
| Amiodarone ($\leq 200\text{mg/day}$) | 797 (68.6%) |
| Dronedarone | 8 (0.7%) |
| Flecainide | 14 (1.2%) |
| Dofetilide | 95 (8.2%) |
| Sotalol | 280 (24.1%) |

**Supplemental Table 4:** Unadjusted perioperative outcomes in recipients with pre-transplant atrial fibrillation, by control agent.

| **Outcome** | **Rate**<br>**(n=1,315)** | **Rhythm**<br>**(n=609)** | **Rate+**<br>**Rhythm**<br>**(n=553)** | **p-value** |
| --- | --- | --- | --- | --- |
| Acute rejection | 265 (20.2%) | 118 (19.4%) | 96 (17.4%) | 0.37 |
| Stroke | 27 (2.1%) | 13 (2.1%) | 18 (3.3%) | 0.27 |
| Dialysis | 161 (12.3%) | 71 (11.7%) | 81 (14.7%) | 0.25 |
| Pacemaker | 38 (2.9%) | 16 (2.6%) | 13 (2.4%) | 0.80 |
| Hospital LOS | 16 (11-23) | 15 (11-24) | 15 (11-22) | 0.58 |
| AF 6m post-tx | 71 (5.8%) | 26 (4.7%) | 39 (7.9%) | 0.09 |
| AF 1y post-tx | 76 (6.9%) | 30 (6.1%) | 45 (9.9%) | 0.053 |

Abbreviations: LOS, length of stay

**Supplemental Table 5:** Adjusted perioperative outcomes in recipients with pre-transplant atrial fibrillation, by control agent.

| **Outcome** | **Adjusted HR (95% CI)** | **p-value** |
| --- | --- | --- |
| Acute rejection (ref: rate-only) |  |  |
| Rhythm-only | 1.34 (0.73-2.47) | 0.34 |
| Rate + rhythm | 0.50 (0.24-1.05) | 0.07 |
| Stroke (ref: rate-only) |  |  |
| Rhythm-only | 1.24 (0.23-6.66) | 0.80 |
| Rate + rhythm | 2.28 (0.53-9.70) | 0.27 |
| Dialysis (ref: rate-only) |  |  |
| Rhythm-only | 0.72 (0.34-1.54) | 0.40 |
| Rate + rhythm | 0.50 (0.22-1.16) | 0.11 |
| Pacemaker (ref: rate-only) |  |  |
| Rhythm-only | 1.08 (0.34-3.39) | 0.90 |
| Rate + rhythm | 0.76 (0.20-2.95) | 0.70 |
| AF 6m post-tx (ref: rate-only) |  |  |
| Rhythm-only | 0.20 (0.02-1.64) | 0.13 |
| Rate + rhythm | 1.84 (0.69-4.93) | 0.22 |
| AF 1y post-tx (ref: rate-only) |  |  |
| Rhythm-only | 0.33 (0.07-1.55) | 0.16 |
| Rate + rhythm | 1.47 (0.58-3.75) | 0.42 |

**Supplemental Table 6:** Risk factors associated with the development of atrial fibrillation in the 6 months post-transplant in those who survive to 6 months.

| Variable | Adjusted Hazards Ratio | 95% Confidence Interval | p-value |
| --- | --- | --- | --- |
| **Donor characteristics** |  |  |  |
| Age (per year) | 0.99 | 0.97-1.02 | 0.60 |
| Male sex | 1.12 | 0.63-1.97 | 0.71 |
| White race | 1.24 | 0.74-2.08 | 0.42 |
| **Recipient characteristics** |  |  |  |
| Age (per year) | 1.00 | 0.98-1.03 | 0.92 |
| Male sex | 0.83 | 0.45-1.53 | 0.56 |
| Race (ref: White) |  |  |  |
| Black | 1.09 | 0.58-2.04 | 0.79 |
| Other | 0.92 | 0.42-2.00 | 0.83 |
| Diagnosis (ref: nonischemic dilated CM) |  |  |  |
| Ischemic CM | 2.08 | 1.17-3.72 | 0.01 |
| Restrictive CM | 1.28 | 0.38-4.37 | 0.69 |
| Hypertrophic CM | 1.94 | 0.64-5.85 | 0.24 |
| Congenital | 0.35 | 0.04-2.92 | 0.33 |
| Other | 0.56 | 0.07-4.24 | 0.58 |
| Pre-transplant IABP | 2.08 | 1.08-4.00 | 0.03 |
| Pre-transplant ECMO | 6.23 | 1.26-30.76 | 0.03 |
| Pre-transplant temporary VAD | 2.46 | 1.29-4.68 | 0.006 |
| Hypertension | 0.80 | 0.49-1.33 | 0.39 |
| Creatinine (per mg/dL) | 1.19 | 0.95-1.50 | 0.13 |
| BMI (per kg/m<sup>2</sup>) | 1.00 | 0.99-1.02 | 0.57 |
| Ischemic time (per hour) | 1.12 | 0.88-1.42 | 0.36 |
| Pre-transplant AF | 2.37 | 1.38-4.09 | 0.002 |

Abbreviations: BMI, body mass index; CM, cardiomyopathy; IABP, intra-aortic balloon pump; VAD, ventricular assist device.

